# Age-structured modelling of Ebola convalescence and re-emergence risk: a Bayesian approach

**DOI:** 10.64898/2026.07.03.26357211

**Authors:** Bruno Enagnon Lokonon, Daniel T. Haydon, Caroline Fakas, Bassirou Bonfoh

## Abstract

**Background:** Increasing evidence indicates that Ebola virus disease (EVD) survivors can remain a source of infection long after clinical recovery. Confirmed survivor-associated transmission events and genomic evidence linking the 2021 Guinea outbreak to viral lineages from the 2013-2016 West African epidemic have demonstrated that persistent infection in survivors can contribute to post-epidemic re-emergence. However, the population-level conditions under which survivor reservoirs may sustain recrudescence remain poorly understood.

**Methods:** We developed an age-structured Bayesian transmission model to quantify survivor-driven recrudescence risk using historical Ebola outbreak data (1976–2022) and empirical viral persistence data from male survivors. Age-specific viral clearance probabilities were estimated for three age groups (≤25, 26–35, and >35 years). The recrudescence reproduction number (*R*_*c*_) was derived using the next-generation matrix approach. Sensitivity analyses examined alternative assumptions regarding viral clearance and the potential contribution of female survivors.

**Results:** The posterior mean recrudescence reproduction number remained below the persistence threshold (*R*_*c*_ = 1) across all viral-clearance scenarios under the assumption of no female survivor contribution. Only by assuming the slowest rate of viral clearance and maximal female survivor contribution did the posterior mean for *R*_*c*_ exceed one (1.052; 95% CrI: 0.428–2.229), suggesting that survivor-driven transmission alone is unlikely to sustain Ebola re-emergence given our current understanding of recrudescence dynamics. Simulations showed that survivor-driven outbreak pressure (rate of survivor-initiated outbreaks) was driven primarily by outbreak size and clustering. Outbreaks involving ≤5,000 EVD cases generally produced outbreak pressure below the estimated natural spillover rate, whereas outbreaks comparable in size to the 2013–2016 West African epidemic generated transient survivor-driven outbreak rates up to 7.8-fold higher than the natural spillover rate before declining to comparable levels within 3–7 years. Moreover, across all viral-clearance scenarios, older (>35 years) male survivors consistently exhibited the longest effective persistence durations and made the largest contribution to the recrudescence reproduction number.

**Conclusions:** The human survivor reservoir represents a plausible complementary pathway for Ebola re-emergence, particularly following large epidemics and should be considered alongside zoonotic spillover as an important source of future outbreaks. Age-dependent viral clearance strongly shapes recrudescence dynamics, with older survivors contributing disproportionately to transmission potential. These findings support age-stratified survivor monitoring, extended persistence surveillance, and improved characterization of viral persistence in both male and female survivors to strengthen post-epidemic preparedness.

## Introduction

Ebola virus disease (EVD) is one of the deadliest infectious diseases known in humans with a mortality rate of 70-90% if untreated (Dyal et al., 2023). Since EVD was first identified in 1976, repeated outbreaks have occurred across Sub-Saharan Africa (Rugarabamu et al., 2020). Recent outbreaks in Uganda and DRC-Uganda border (2025-2026) underscore the critical need to understand diverse disease transmission pathways, including survivor-driven recrudescence (European CDC, 2026). For decades, Ebola epidemics were viewed as independent zoonotic events (Fairhead et al., 2021). Spillover from wildlife reservoirs, possibly fruit bats (Abah et al., 2024), introduced the virus into human populations, after which chains of human-to-human transmission were eventually interrupted through isolation, contact tracing, and public health control measures. This view, although useful, is now incomplete (Fairhead et al., 2021).

The 2013-2016 West African epidemic profoundly changed the understanding of EVD transmission and persistence. The epidemic caused more than 28,000 reported cases and over 11,000 deaths across Guinea, Sierra Leone, and Liberia (Thorson et al., 2021; Bosa et al., 2024). It was the largest Ebola outbreak ever recorded. More importantly, it generated, for the first time, a survivor population large enough to systematically study the phenomenon. Evidence emerging from the West Arican’s cohort demonstrated that EVD can persist in individuals long after clinical recovery (Fairhead et al., 2021). We describe these individuals as convalescent.

After acute infection, EVD may persist within immune-privileged sites where immune surveillance is reduced, including the testes, eyes, and central nervous system (Meek et al., 2025). In male survivors, the testes appear to function as a long-term viral reservoir (Thorson et al., 2021). Viral RNA can remain detectable in semen during asymptomatic convalescence for months or even years after recovery. Deen et al. (2017) detected EVD RNA in the semen of 27% of male survivors in Sierra Leone. Fischer et al. (2017) reported persistence beyond two years after acute illness. In Liberia, one survivor remained semen-positive 988 days after hospital discharge (Meek et al., 2025). These findings are no longer considered isolated events. They suggest that a prolonged convalescent state from which virus may recrudesce may represent an important feature of EVD biology.

The epidemiological implications are substantial. In 2015, sexual transmission from a male survivor was confirmed in Liberia (Christie et al., 2015; Mate et al., 2015). Several additional probable sexual transmission events were subsequently documented. Soka et al. (2016), analysing Liberia’s national semen testing programme, reported that 9% of 429 enrolled male survivors produced at least one positive semen sample, with persistence detected up to 565 days after discharge. In Guinea, a survivor-associated transmission chain produced 13 secondary cases 531 days after the survivor’s original illness (Diallo et al., 2016). The strongest evidence emerged in 2021, when genomic sequencing linked a new outbreak in Guinea to viral lineages circulating during the 2013-2016 epidemic, indicating recrudescence from a survivor more than five years after the initial infection (Keita et al., 2021). These observations established that Ebola outbreaks may re-emerge not only from wildlife reservoirs, but also from convalescent human survivors (Meek et al., 2025).

The duration of the convalescent state is not uniform across survivors. Age is one of the strongest predictors of viral clearance duration. Soka et al. (2016) first noted that men older than 40 years were disproportionately represented among semen-positive survivors. Thorson et al. (2021), using a prospective cohort of 220 male survivors in Sierra Leone, showed that men aged 25 years or younger were 3.17 times more likely to clear EVD RNA from semen than men older than 35 years. Men aged 26-35 years showed intermediate clearance dynamics. Among older survivors with severe disease, persistence exceeded 20% one year after discharge (Thorson et al., 2021). Dyal et al. (2023) later confirmed the age effect in a Liberian cohort and identified additional correlates of delayed clearance, including elevated serum IgG3 levels, HLA-C*03:04 expression, and ocular complications. Together, these findings suggest that age-related immune mechanisms may strongly influence long-term EVD persistence.

The contribution of female survivors to post-epidemic transmission is uncertain. Large longitudinal datasets on female genital shedding do not currently exist. Nevertheless, EVD RNA has been detected in vaginal secretions, breast milk, and other bodily fluids (Keita et al., 2019; Meek et al., 2025). Female-to-male transmission therefore cannot be excluded. Any quantitative framework that omits female survivors from the convalescent reservoir is likely to underestimate uncertainty in survivor-driven transmission.

Despite growing evidence for survivor-associated recrudescence, most mathematical models still treat survivors as homogeneous populations (Vinson et al., 2016; Abbate et al., 2016). Our overall objective was to provide the first age-stratified quantitative assessment of survivor-driven EVD re-emergence risk and to identify implications for post-epidemic surveillance and survivor monitoring strategies. Our study was guided by three main research questions. First, can survivor-driven EVD transmission alone sustain epidemic re-emergence under biologically plausible persistence conditions? Second, how sensitive are recrudescence dynamics to assumptions regarding the duration of the convalescent state? Third, how strongly does age-dependent viral clearance influence the contribution of survivors to recrudescence reproduction number?

## 2. Methods

### 2.1 Model formulation

We developed a stochastic age-structured transmission model describing the accumulation and persistence of male Ebola convalescent survivors. The male convalescent population in age group *g* at time *t*+1 is the sum of: (i) surviving convalescents from time *t* who have not cleared the virus, (ii) new male survivors from Ebola cases occurring in (*t, t*+1) allocated proportionally to age group *g*, and (iii) net aging flows between adjacent age groups. Using sex-specific Ebola mortality data from Liberia reported by Matson et al. (2022), 50% of survivors were male (*p*_*m*_= 0.5). The model is given by:

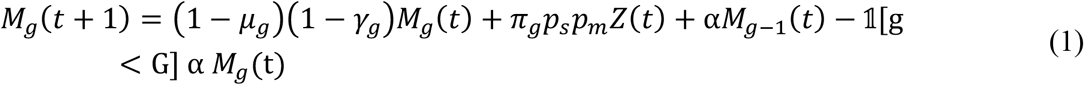

where the time units are assumed to be years and:

*M*_*g*_(*t* + 1) is the male convalescent population in age group *g* at time *t*+1;

*M*_*g*_(*t*) is the male convalescent population in age group *g* at time *t*;

*p*_*s*_ is the annual survival probability from Ebola disease;

*p*_*m*_ is the proportion of survivors who are male;

*π*_*g*_ is the proportion of male survivors belonging to age group *g*;

*μ*_*g*_ is the age-specific annual natural death probability;

*γ*_*g*_ is the age-specific annual viral clearance probability (after which convalescents are no longer able to transmit);

*Z*(*t*) is the total number of new Ebola cases between time t and t+1;

α is the annual aging-out rate from one age band to the next;

*G* denotes the total number of age groups considered in the model.

Boundary conditions: *M*_*g*−1_(*t*) = 0 for *g* = 1 (no inflow to youngest group), α *M*_*g*_(*t*) = 0 for *g* = *G* (no aging out of oldest group), [g < *G*] indicator is equal to 1 if not in oldest group, 0 otherwise.

Aging transitions were found to have negligible effects on model outputs and were therefore excluded from the final model for parsimony. The resulting simplified formulation is:

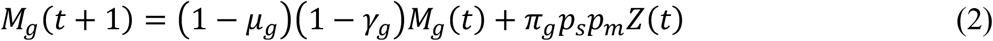

Equation (2) constitutes the model implemented and used for all subsequent analyses.

### 2.2 Age structure and clearance rate derivation

We stratified the male convalescent population into *G* = 3 age groups: <25, 26–35, and 35+ years. Age-group proportions (*π*_*g*_) were derived from a study by Thorson et al. (2021), which analysed semen persistence in one of the largest cohorts of male Ebola survivors (*n* = 219). The proportion of male survivors in each age group was calculated as:

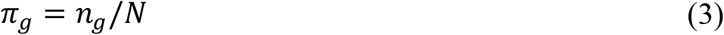

where *n*_*g*_ is the number of individuals in age group *g* and *N* is the total sample size.

Younger men (≤25 years), middle age men (26–35 years), and older men (>35 years), represented 30.6 %, 41.6 %, and 27.8 %, respectively. This structure was selected because Thorson et al. (2021) demonstrated a clear age-dependent pattern in viral clearance, with younger men (≤25 years) being 3.17 times more likely and men aged 26–35 years being 1.85 times more likely to become EVD RNA negative than men aged >35 years. The cohort demographics were also comparable to the national male survivor registry (Thorson et al., 2021), supporting its use as a baseline representation of the survivor population. *μ*_*g*_ was set equal to 0.006, 0.009, 0.017 per convalescent per year for younger men (≤25 years), middle-age men (26-35 years), and older men, respectively (LISGIS, 2022).

Let *D*_*g*_ denote the mean number of days of viral detection in semen in age group *g*. To analyse a range of recovery scenarios while preserving the empirical age-specific pattern, we defined a relative clearance multiplier:

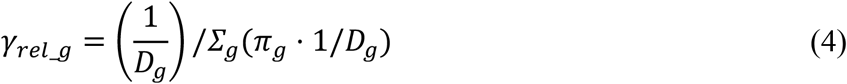

For each scenario with overall recovery probability *ε*_*m*_ (average annual clearance probability), age-specific rates are then:

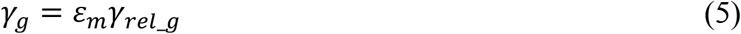

The semen dataset, comprising 59 observations, was used to estimate age-specific persistence durations (*D*_*g*_) and the relative viral clearance multiplier. However, available persistence studies are characterized by limited sample sizes, intermittent sampling schedules, and relatively short follow-up periods (Meek et al. 2025). Consequently, substantial uncertainty remains regarding the true population-level viral clearance rate. To quantify this uncertainty, we fitted an exponential distribution to the observed persistence durations. The estimated average annual clearance probability was *ϵ*_*m*_ = 0.68, with approximate 95% confidence interval bounds of 0.57 and 0.77. Additional scenarios (0.22, 0.42, and 0.83) were obtained by varying the fitted exponential rate parameter by multiples of two standard errors and converting the resulting rates to annual clearance probabilities. We therefore considered six clearance scenarios:

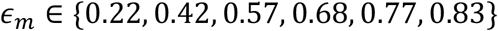

### 2.3 Outbreak and case process

The number of Ebola outbreaks occurring in year *t* was assumed to follow a Poisson distribution:

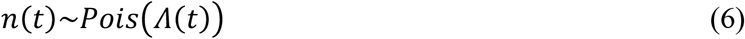

where the annual outbreak rate is:

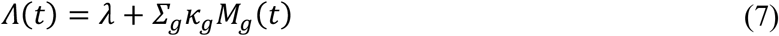

*λ* represents the annual rate of zoonotic spillover events leading to detected onward transmission in humans, while *κ*_*g*_ is the per-capita recrudescence transmission rate associated with convalescent survivors in age group *g*, expressed per convalescent per year.

Conditional on *n*(*t*) outbreaks occurring in year *t*, the total annual number of Ebola cases *Z*(*t*) was assumed to follow a Negative Binomial distribution:

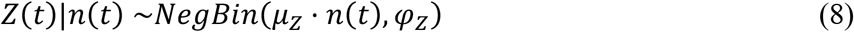

where *μ*_*Z*_ is the mean number of Ebola cases generated per outbreak and *φ*_*Z*_ is an overdispersion parameter capturing variability in annual outbreak-associated case totals. Both parameters were estimated by fitting the model to observed annual Ebola case totals conditional on outbreak occurrence. The estimations yielded a mean outbreak size of *μ*_*Z*_ = 970.61 and an overdispersion parameter of *φ*_*Z*_ = 0.22.

### 2.4 Prior distributions and Bayesian inference

We assigned prior distributions to the model parameters estimated by Markov Chain Monte Carlo (MCMC). The priors and their justifications are in Table 1:

**Table 1:**
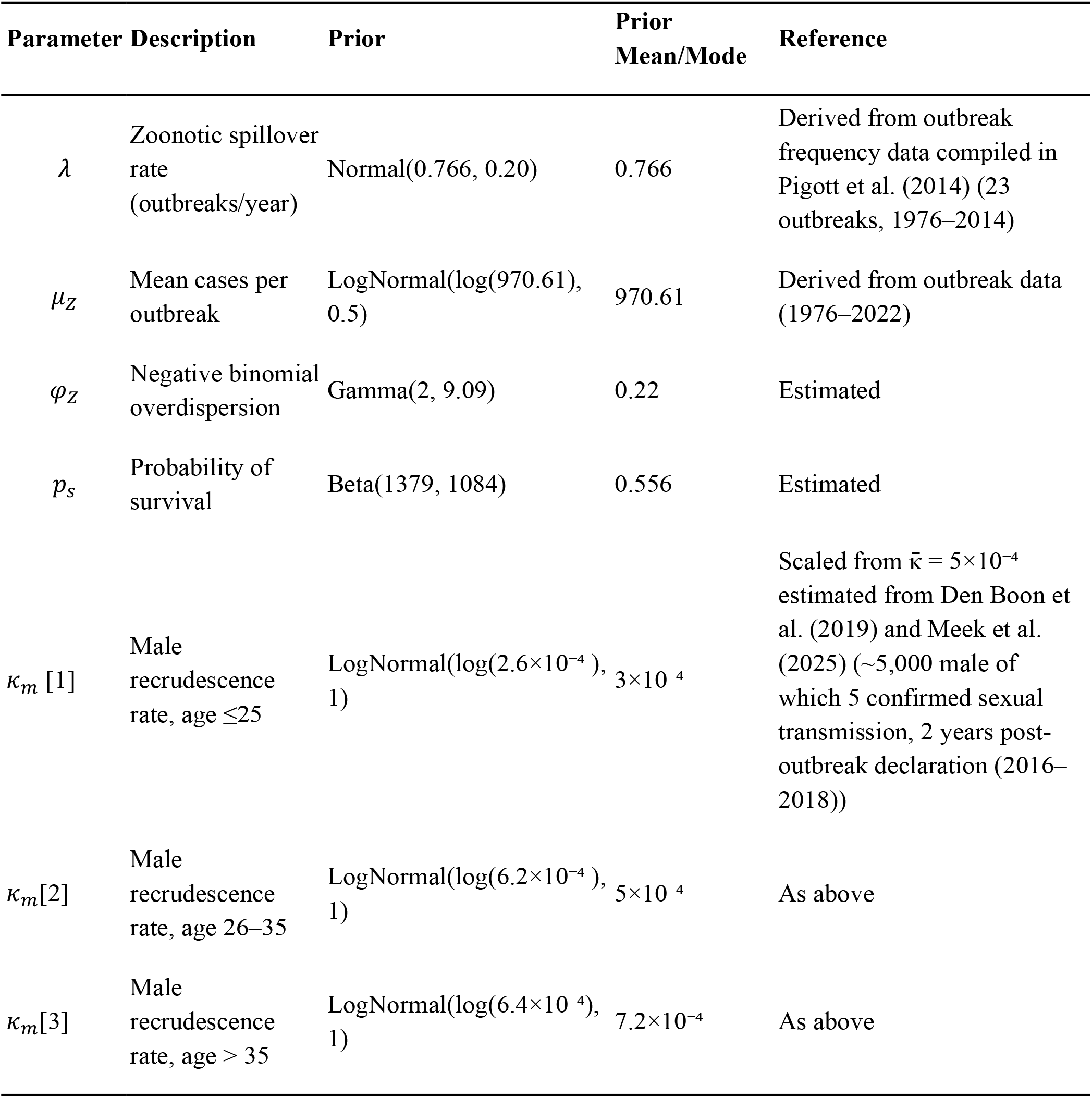
Values of priors used in the model (2)

The fixed parameters are presented in Table 2.

**Table 2:** Fixed parameters used in the model (2)

| Parameter | Description | Value(s) | Source |
| --- | --- | --- | --- |
| $p_m$ | Proportion of Ebola survivors assumed to be male | 0.50 | Matson et al. (2022) |
| $\pi_g$ | Proportion of male survivors by age group | $\leq 25$: 0.306; 26–35: 0.416; $> 35$: 0.278 | Thorson et al. (2021) |
| $D_g$ | Mean duration of EBOV RNA detection in semen by age group | $\leq 25$: 121.43 days; 26–35: 198.74 days; $> 35$: 457.79 days | Semen persistence dataset |
| $\gamma_{rel\_g}$ | Relative age-specific viral clearance multiplier | $\leq 25$: 1.58; 26–35: 0.96; $> 35$: 0.42 | Derived from $D_g$ |
| $\mu_g$ | Age-specific annual natural mortality probability | $\leq 25$: 0.006; 26–35: 0.009; $> 35$: 0.017 | LISGIS (2022) |
| $\epsilon_m$ | Population-average male viral clearance probability (prior means) | 0.22, 0.42, 0.57, 0.68, 0.77, 0.83 | Scenario assumptions |

We fitted the model using Bayesian inference implemented in Stan (Annis et al., 2017), running four Markov chains with 4,000 iterations each and discarding the first 2,000 iterations of each chain as warm-up. Convergence was assessed using the Gelman–Rubin statistic (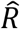), effective sample size (ESS), and visual inspection of trace plots. Across all analyses, no divergent transitions were observed, no chains reached the maximum tree depth, and E-BFMI diagnostics indicated satisfactory Hamiltonian Monte Carlo performance. All monitored parameters showed satisfactory convergence.

### 2.5 Endemic equilibrium

At the endemic equilibrium, 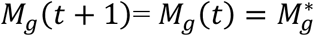 for all *g*. Defining the mean outbreak size as *μ*_*Z*_ and the mean number of outbreaks as Λ^∗^ at this endemic equilibrium, from Eq. 2:

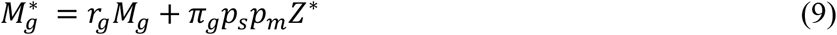

with *r*_*g*_ = (1 − *μ*_*g*_)(1 − *γ*_*g*_), and:

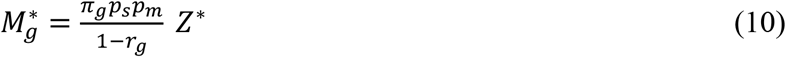

The expected number of cases per year at endemic equilibrium is

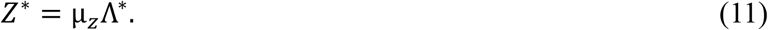

Then:

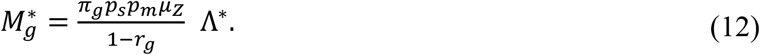

Let:

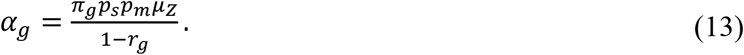

Then:

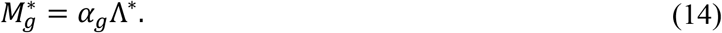

Now Λ^∗^ becomes:

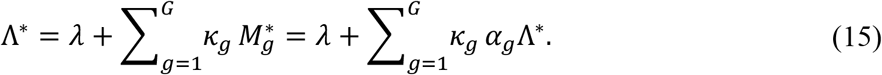

Defining:

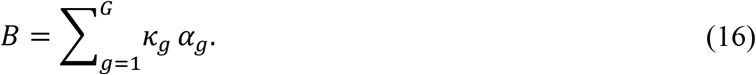

Then:

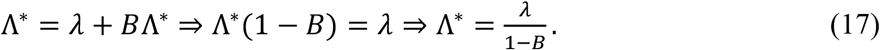

Finally:

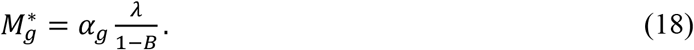

A positive finite endemic equilibrium exists if and only if *B* < 1.

### 2.6 Recrudescence reproduction number(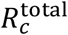)

We term 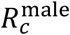 the reservoir persistence threshold, and define it as the expected number of new male convalescents generated by introducing a single male convalescent into an otherwise disease-free male population (*M*_*g*_= 0 for all *g*), using the next-generation matrix approach (Diekmann et al., 2010). The next-generation matrix K has elements:

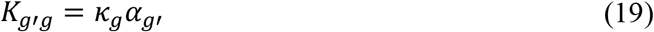

Since K = ακ^T^ is a rank-1 matrix, its spectral radius, the recrudescence reproduction number, simplifies to:

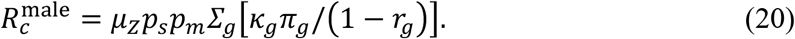

This is identical to the stability parameter *B*, implying that 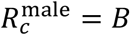.

Because longitudinal data on viral persistence among female survivors are currently very limited to parameterize a separate transmission model, female contribution was incorporated through a scaling parameter, *θ*_*f*_, representing the contribution of female survivors relative to male survivors. The total recrudescence reproduction number was therefore defined as:

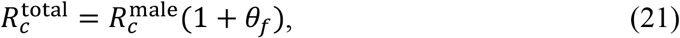

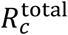 is then the threshold parameter governing survivor-driven transmission:

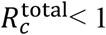: survivor-driven transmission is self-limiting and the reservoir remains bounded;

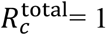: the system is at the human reservoir persistence threshold;

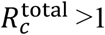: survivor-driven transmission can sustain itself and generate a growing reservoir under the deterministic mean-field approximation (i.e., in the absence of limiting mechanisms).

### 2.7 Sensitivity analysis

Sensitivity analyses were conducted to evaluate uncertainty in both viral clearance and female survivor contribution to transmission. Viral-clearance uncertainty was explored through alternative values of the average annual clearance probability (*ϵ*_*m*_=0.22, 0.42, 0.57, 0.68, 0.77, 0.83), while uncertainty in female transmission was represented using a scaling factor (*θ*_*f*_) relative to the estimated male survivor contribution. Female contribution was parameterized using fixed scaling factors *θ*_*f*_ ∈ {0, 0.25, 0.5, 1.0}, corresponding to no female contribution, 25%, 50%, and 100% of the estimated male survivor contribution to transmission, respectively.

To illustrate the epidemiological implications of uncertainty in the male viral clearance probability, we performed simulations of survivor-driven outbreak pressure following one, two, or three clustered Ebola outbreaks. Outbreak sizes of 1,000, 5,000, 10,000 and 30,000 total EVD cases were considered. For each scenario, newly generated male survivors entered the survivor reservoir according to the estimated survivor proportion, and the reservoir subsequently declined through viral clearance. Survivor-driven outbreak pressure (rate of survivor-initiated outbreaks) was calculated from the male survivor reservoir and compared with the estimated natural spillover rate (*λ* = 0.675 outbreaks per year). The baseline analysis used *ϵ*_*m*_= 0.68, while uncertainty was illustrated using *ϵ*_*m*_ = 0.57-0.77.

Finally, we examined how alternative viral clearance scenarios influenced age-specific persistence by calculating the effective persistence duration (365/*γ*_*g*_) for each age group. These analyses were used to quantify the relative contribution of younger (≤25 years), middle-aged (26–35 years), and older (>35 years) male survivors to the survivor reservoir and the male recrudescence reproduction number.

## 3. Results

### 3.1 Posterior parameter estimates and model calibration

Figure 1 presents the prior distributions and posterior density estimates for the seven fitted model parameters. The 95% posterior credible intervals are shown by the grey shaded regions. Posterior inference yielded mean estimates of the annual spillover outbreak rate *λ* = 0.675 outbreaks per year (95% CrI: [0.472, 0.901]), the mean outbreak size *μ*_*Z*_= 991.76 cases per outbreak (95% CrI: [494.43, 1853.33]), the outbreak-size dispersion parameter *φ*_*Z*_= 0.243 (95% CrI: [0.146, 0.364]), and the Ebola survival probability *p*_*s*_= 0.559 (95% CrI: [0.540, 0.579]). The age-specific recrudescence transmission rates were estimated as *κ*_*m*_[1] = 2.52e-04 per younger men per year (95% CrI: [9.60e-05, 5.35e-04]), *κ*_*m*_[2] = 3.18e-04 per middle age men (95% CrI: [1.30e-04, 6.23e-04]), and *κ*_*m*_[3]=4.64e-04 per older men (95% CrI: [1.96e-04, 8.85e-04]). Posterior distributions were generally narrower than their corresponding priors, indicating that the data reduced uncertainty in the model parameters. The strongest posterior updating was observed for the age-specific recrudescence rates (*κ*_*m*_[1], *κ*_*m*_[2], and *κ*_*m*_[3]), whereas *p*_*s*_ remained close to its prior expectation.

**Figure 1.**
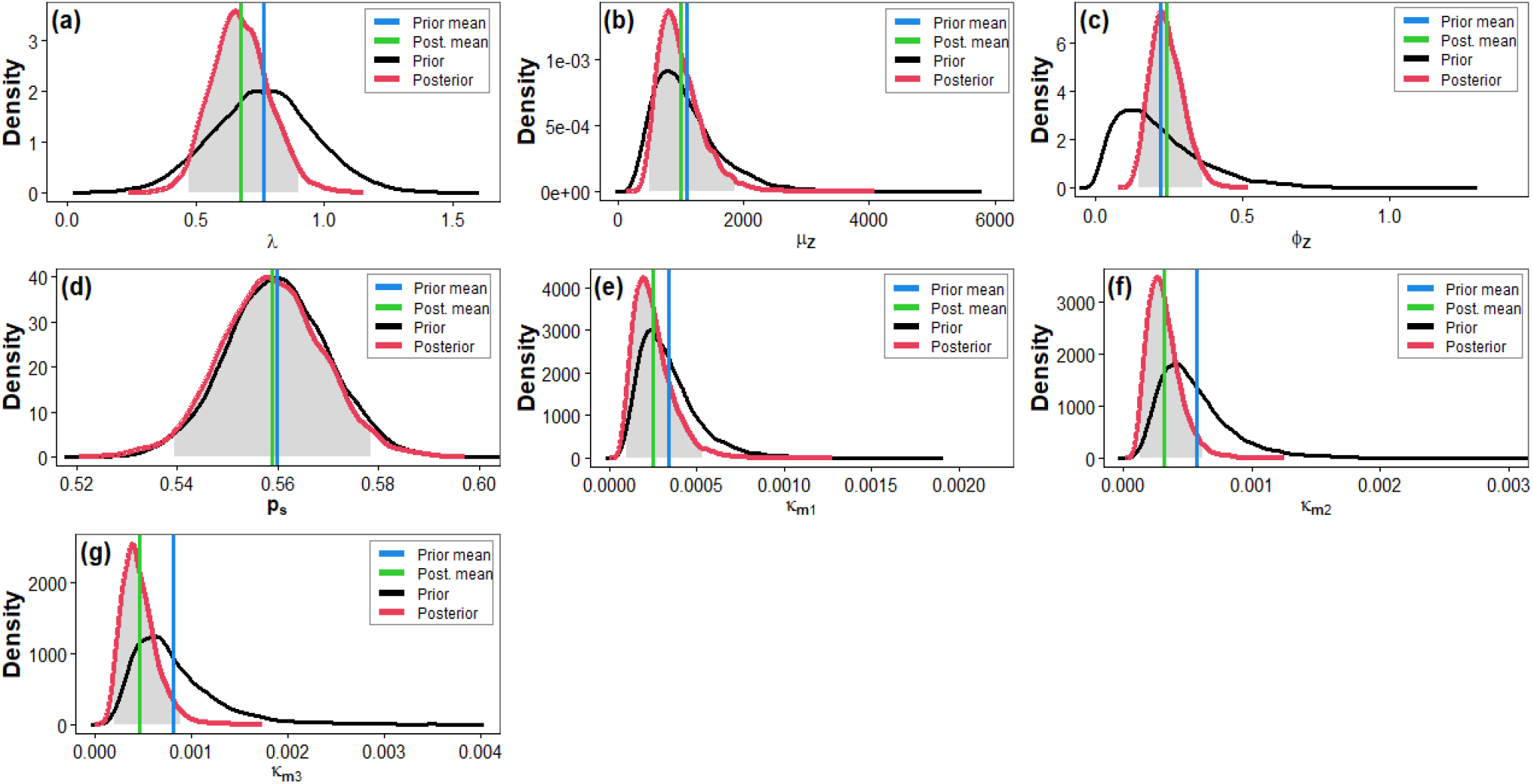
Prior and posterior distributions of the seven core model parameters. Black curves represent prior distributions and red curves posterior distributions. Blue and green vertical lines indicate prior and posterior means, respectively. Grey shading denotes the 95% posterior credible interval.

### 3.2 Recrudescence reproduction number under alternative clearance assumptions and female contribution

Under the assumption of no female survivor contribution (*θ*_*f*_= 0), the estimated total recrudescence reproduction number (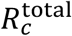) decreased substantially as the male viral clearance probability (*ϵ*_*m*_) increased (Figure 2). Posterior mean estimates declined from 0.526 (95% CrI: 0.214–1.115) at *ϵ*_*m*_= 0.22 to 0.193 (95% CrI: 0.075–0.421) at *ϵ*_*m*_= 0.83. Although posterior means remained below the persistence threshold (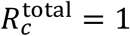) across all scenarios, the upper credible interval exceeded one for the slowest clearance scenario (*ϵ*_*m*_= 0.22). This indicates that slow viral clearance increases the potential for sustained survivor-driven transmission.

**Figure 2.**
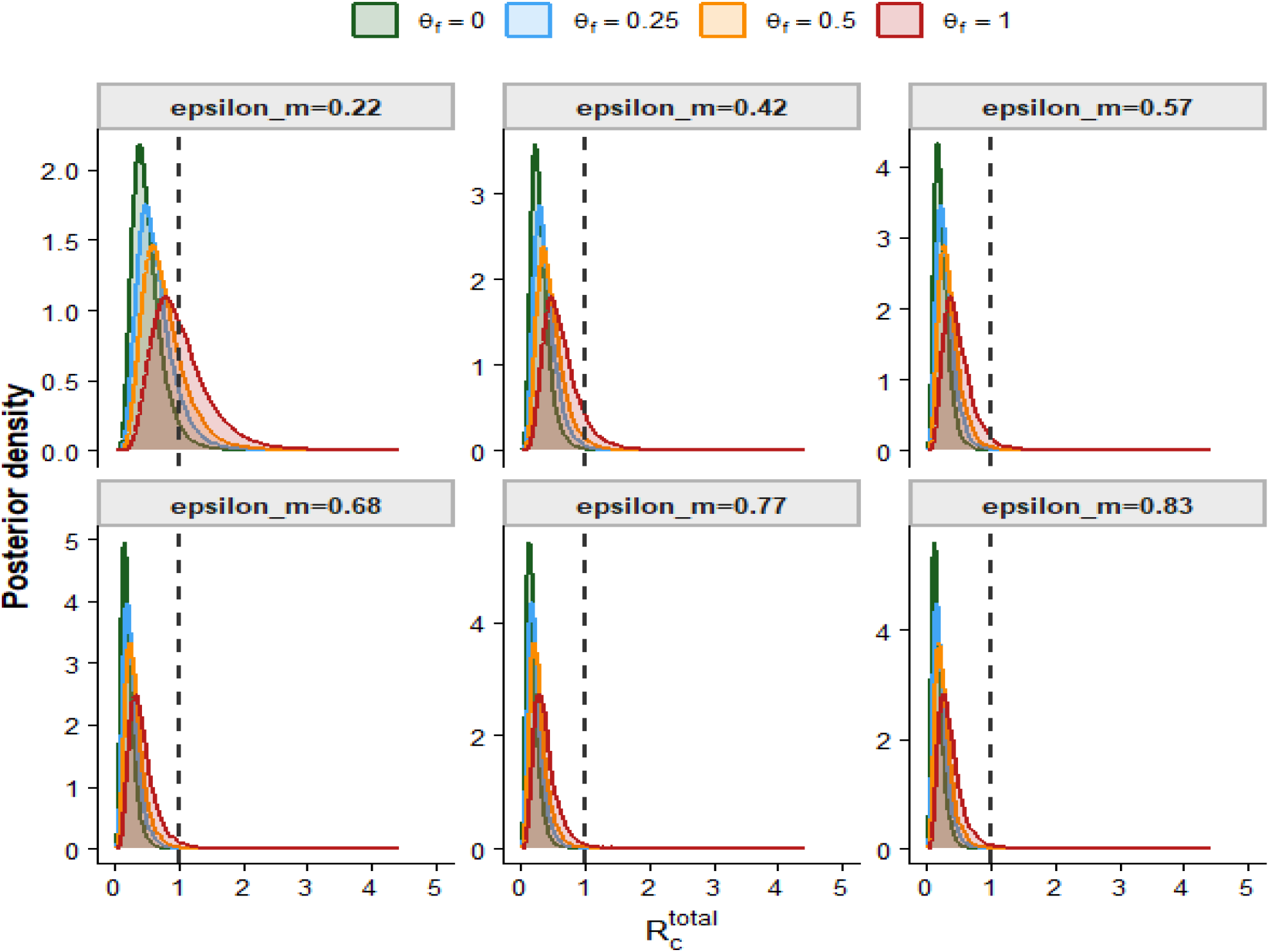
Posterior distributions of the total recrudescence reproduction number 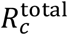 under the viral clearance scenarios. Panels correspond to the priors for the male viral clearance probability (*ϵ*_*m*_). Colours indicate alternative female contribution assumptions (*θ*_*f*_ = 0, 0.25, 0.5, and 1). Vertical dashed lines denote 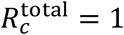.

Increasing female survivor contribution consistently increased transmission potential across all clearance scenarios. Under the slowest clearance scenario (*ϵ*_*m*_= 0.22), the posterior mean 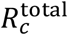 increased from 0.526 (95% CrI: 0.214–1.115) when *θ*_*f*_= 0 to 1.052 (95% CrI: 0.428–2.229) under maximal female contribution (*θ*_*f*_= 1), with the posterior probability that 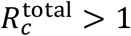 increasing from 4.2% to 45.8%. For intermediate clearance rates (*ϵ*_*m*_= 0.42–0.57), female contribution increased posterior means but only the upper credible intervals approached or slightly exceeded one. Under faster clearance scenarios (*ϵ*_*m*_ ≥ 0.68), posterior means and credible intervals remained below the persistence threshold even under maximal female contribution.

### 3.3. Transient survivor-driven outbreak pressure following clustered Ebola outbreaks

Simulations showed that short-term survivor-driven outbreak pressure (i.e., the expected rate of survivor-initiated outbreaks) was primarily determined by outbreak size and the number of closely clustered outbreaks (Figure 3). Uncertainty in the male viral clearance probability within the baseline range (*ϵ*_*m*_=0.57-0.77) had only a modest effect.

**Figure 3.**
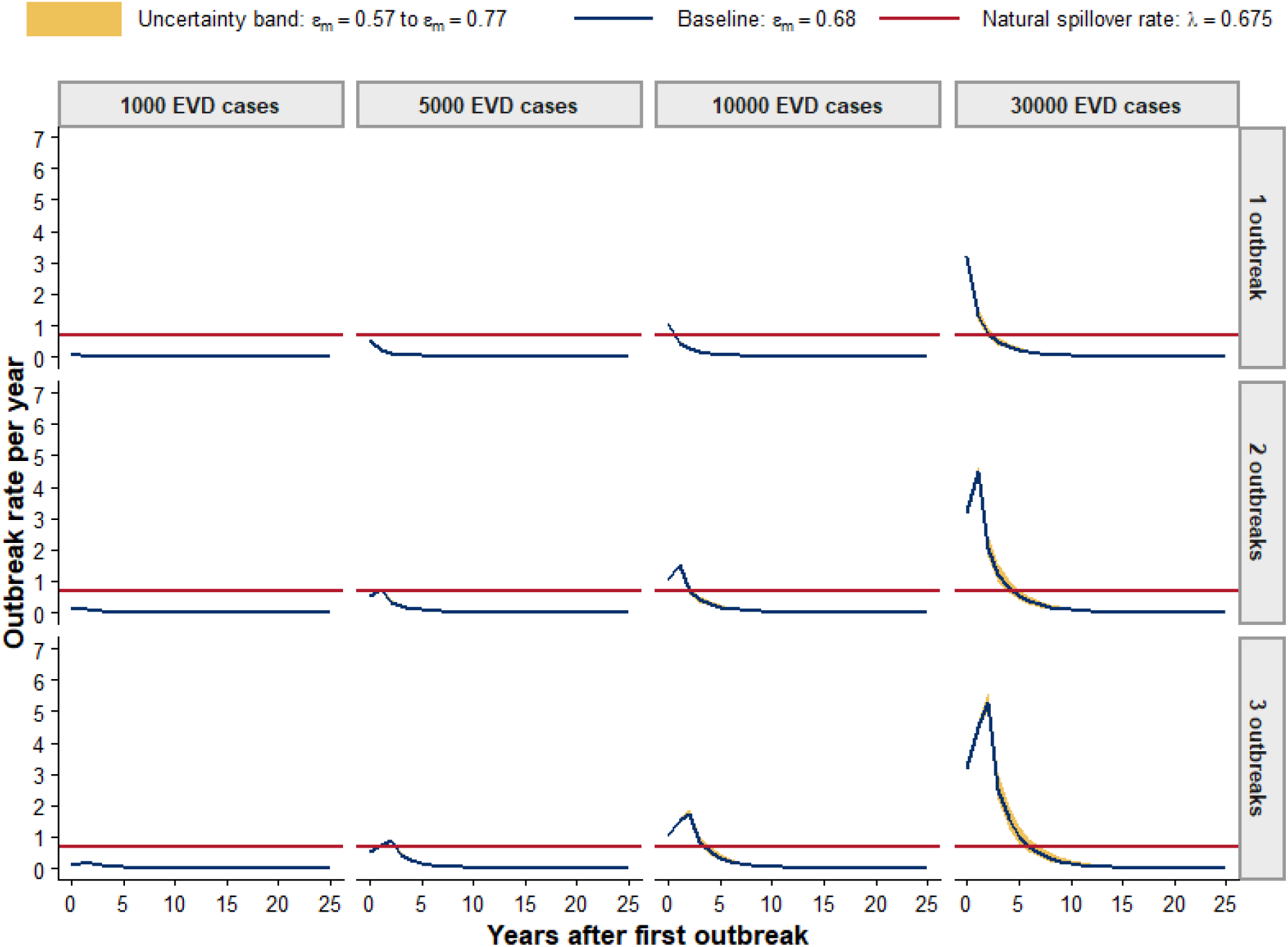
Deterministic survivor-driven outbreak pressure following clustered Ebola outbreaks. Blue lines show the baseline scenario (*ϵ*_*m*_=0.68), the shaded region represents uncertainty (*ϵ*_*m*_=0.57-0.77), and the red horizontal line indicates the estimated natural spillover rate (*λ*= 0.675 outbreaks per year).

For 1,000 EVD cases, the expected rate of survivor-initiated outbreaks remained well below the estimated natural spillover rate in all scenarios. The maximum outbreak pressure reached only 26% of the natural spillover rate, even after three clustered outbreaks. With 5,000 EVD cases, the expected rate of survivor-initiated outbreaks increased but exceeded the natural spillover rate only after two or three clustered outbreaks. The peak of the expected rate of survivor-initiated outbreaks reached 1.1 and 1.3 times the natural spillover rate, respectively, and remained above this threshold for only 1-2 years. During this period, outbreaks originating from the survivor reservoir were predicted to occur more frequently than zoonotic spillover events. For 10,000 EVD cases, the expected rate of survivor-initiated outbreaks exceeded the natural spillover rate even after a single outbreak. The peak of the expected rate of survivor-initiated outbreaks reached approximately 1.6, 2.2, and 2.6 times the natural spillover rate following one, two, and three clustered outbreaks, respectively. The rate of survivor-initiated outbreaks remained above the natural spillover rate for 1-4 years before declining as the survivor reservoir contracted through viral clearance.

Under a West Africa epidemic-sized scenario (30,000 EVD cases), the rate of survivor-initiated outbreaks increased substantially. The peak of this rate reached approximately 4.8, 6.7, and 7.8 times the natural spillover rate following one, two, and three clustered outbreaks, respectively. This indicates that, for several years after a very large epidemic, survivor-driven transmission could temporarily become the dominant source of new outbreaks. However, transmission pressure declined rapidly and returned to the level of the natural spillover rate within 3-7 years as the survivor reservoir progressively cleared infection.

### 3.3 Age-specific contributions to recrudescence

Age-specific contributions to the male recrudescence reproduction number (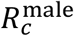) differed consistently across survivor age groups (Figure 4). Older male survivors (>35 years) contributed the largest share of 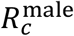 across all viral-clearance scenarios, followed by men aged 26–35 years. The absolute contribution of each age group declined as the viral clearance probability increased.

**Figure 4.**
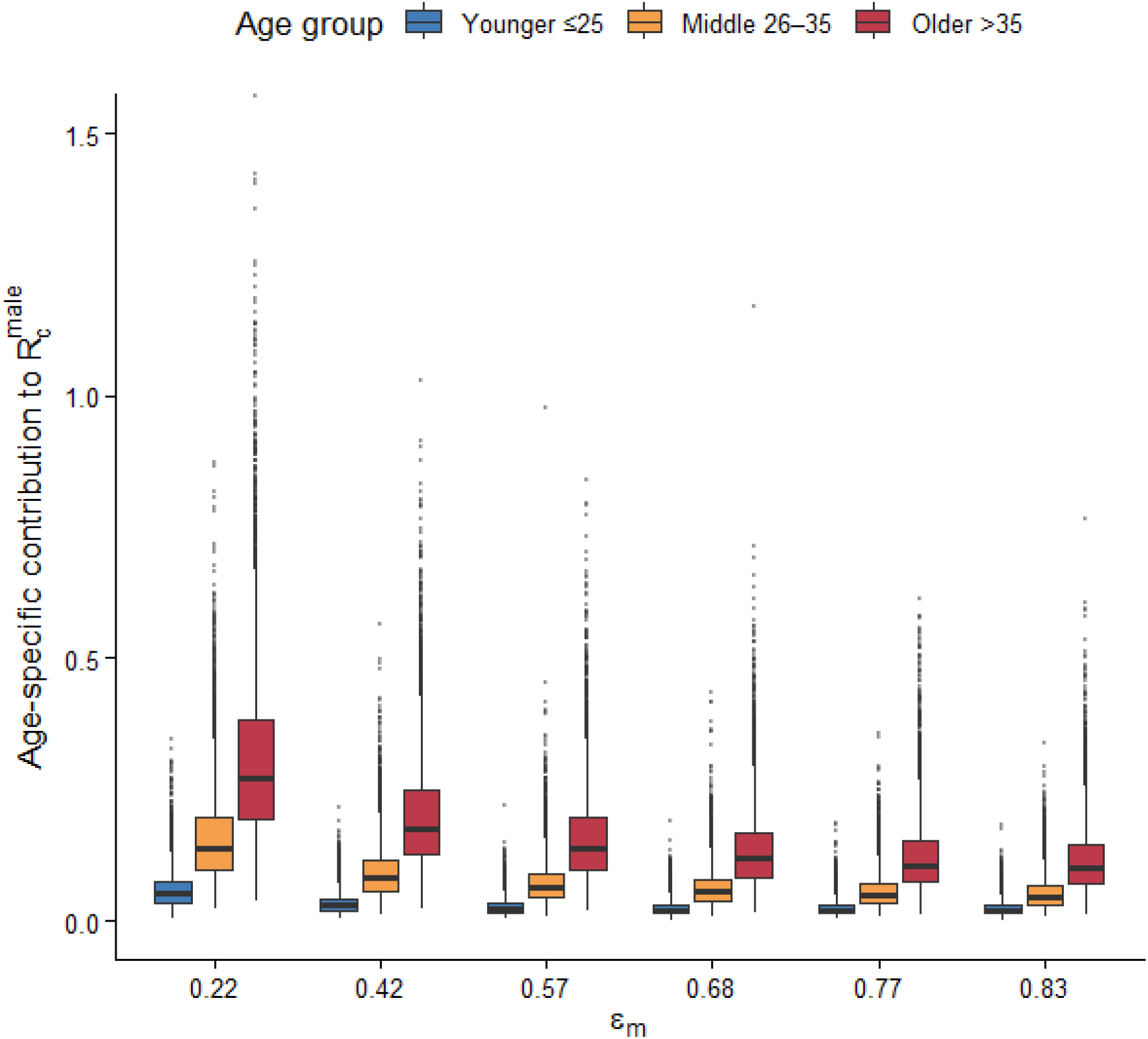
Age-specific contributions to the male recrudescence reproduction number under alternative viral clearance assumptions

Under the slowest clearance scenario (*ϵ*_*m*_= 0.22), older male survivors contributed a posterior mean of 0.308 (95% CrI: 0.097–0.727), compared with 0.159 (95% CrI: 0.049–0.387) among men aged 26–35 years and 0.059 (95% CrI: 0.017–0.153) among younger men (≤25 years). Thus, older men contributed approximately 1.9 times more than middle-aged men and 5.2 times more than younger men, whereas middle-aged men contributed about 2.7 times more than younger men. The same ranking persisted across all other viral-clearance scenarios.

### 3.4 Effective persistence duration across age groups and clearance scenarios

The effective persistence duration of the male survivor reservoir declined markedly as the viral clearance probability increased (Figure 5). The largest reduction occurred between *ϵ*_*m*_= 0.22 and 0.57, after which persistence durations decreased more gradually, particularly for younger survivors. Across all clearance scenarios, a consistent age gradient was observed: older survivors (>35 years) exhibited the longest persistence durations, followed by middle-aged survivors (26– 35 years) and younger survivors (≤25 years).

**Figure 5.**
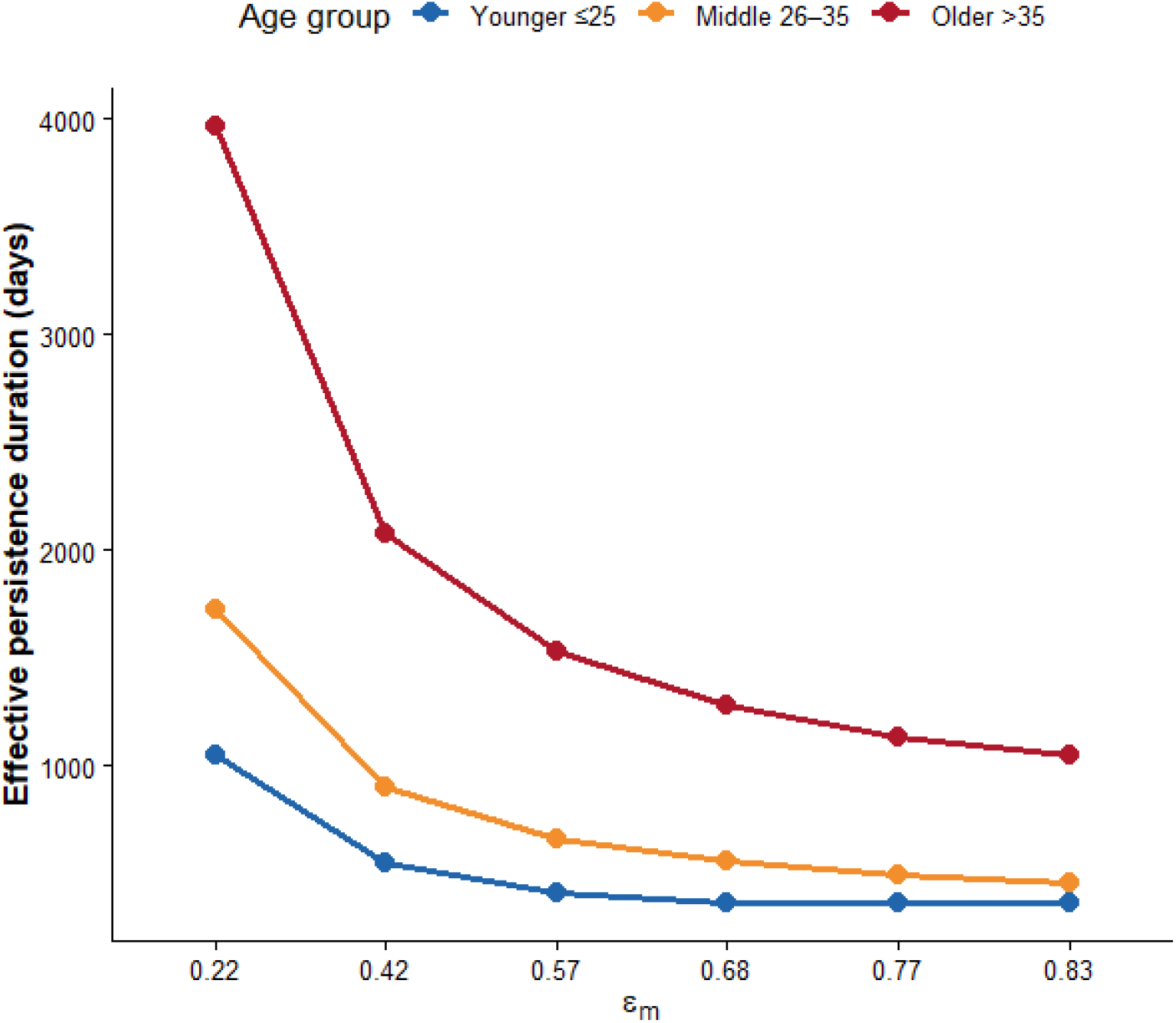
Effective persistence duration across age groups, viral-clearance scenarios, and persistence assumptions.

Under the slowest clearance scenario (*ϵ*_*m*_=0.22), the estimated persistence duration was 3,964 days for older survivors, 1,721 days for middle-aged survivors, and 1,051 days for younger survivors. Older survivors therefore persisted approximately 2.3 times longer than middle-aged survivors and 3.8 times longer than younger survivors.

As viral clearance increased, persistence durations decreased across all age groups. Under the baseline scenario (*ϵ*_*m*_=0.68), estimated persistence durations declined to 1,282, 557, and 365 days for older, middle-aged, and younger survivors, respectively. At the highest clearance scenario (*ϵ*_*m*_=0.83), persistence durations further declined to 1,051, 456, and 365 days.

## 4. Discussion

### 4.1 Survivor-driven recrudescence as a quantifiable epidemiological pathway

The recognition that Ebola recrudescence can originate from persistent human infection, rather than exclusively from wildlife spillover, represents an important shift in post-epidemic risk assessment. Several documented transmission events have demonstrated that survivors may remain a source of infection long after clinical recovery. Sexual transmission in Liberia and survivor-associated transmission chains in Guinea showed that convalescent shedding can continue far beyond conventional monitoring periods (Christie et al., 2015; Mate et al., 2015; Diallo et al., 2016). More recently, genomic investigations linked the 2021 Guinea outbreak to viral lineages circulating during the 2013–2016 West African epidemic, providing compelling evidence that persistent infection in a survivor can reintroduce transmission years after an outbreak has apparently ended (Keita et al., 2021).

While these observations established the biological plausibility of survivor-driven recrudescence, they did not quantify the epidemiological conditions under which such events could sustain transmission. Our model addresses this gap by estimating the recrudescence reproduction number (*R*_*c*_) directly from empirical persistence data. Across most scenarios, the posterior mean reproduction number remained below the persistence threshold. However, under the slowest viral-clearance scenario, the upper credible interval exceeded one, and increasing female contribution further increased the probability that survivor-driven transmission could become self-sustaining. These results indicate that the survivor reservoir is unlikely to sustain transmission under typical clearance assumptions but may become epidemiologically important when viral clearance is substantially delayed.

The outbreak simulations further illustrate the epidemiological consequences of the survivor reservoir. Following relatively small outbreaks (≤5,000 EVD cases), survivor-driven outbreak pressure generally remained below the estimated natural spillover rate. In contrast, outbreaks comparable in size to the 2013–2016 West African epidemic generated a transient period during which survivor-driven outbreaks were predicted to occur more frequently than zoonotic spillover events. This elevated transmission pressure declined over subsequent years as infected survivors progressively cleared infection. Together, these results suggest that the epidemiological importance of the survivor reservoir depends not only on viral clearance but also on the scale and clustering of preceding epidemics. The survivor reservoir should therefore be considered a complementary source of Ebola re-emergence alongside wildlife spillover, particularly following large epidemics.

### 4.2 Viral clearance remains a key source of uncertainty in survivor-driven transmission

The sensitivity analyses identified viral clearance as one of the principal determinants of survivor-driven transmission potential. Although the estimated recrudescence reproduction number declined steadily as the annual viral clearance probability increased, viral clearance remained a major source of uncertainty in the model predictions. These findings highlight the need for more accurate estimates of viral clearance dynamics among Ebola survivors.

Current evidence on viral persistence remains incomplete. A systematic review by Cheung et al. (2019) estimated a mean persistence duration of approximately one year among male survivors. However, prospective cohort studies have continued to detect Ebola virus RNA at the end of follow-up, suggesting that persistence durations may be underestimated because of right-censoring and limited observation periods (Meek et al., 2025). Persistence beyond two years has also been documented in survivor cohorts (Fischer et al., 2017), demonstrating substantial heterogeneity in viral clearance.

An important observation is that the geometric clearance process assumed in our model reproduced the prolonged persistence tail reported by Meek et al. (2025) reasonably well. This supports the use of a constant annual viral clearance probability as a parsimonious representation of survivor recovery while allowing a small proportion of survivors to remain infectious for extended periods. Nevertheless, more detailed longitudinal data are needed to determine whether more flexible clearance functions better capture individual variation in viral persistence.

Overall, these findings suggest that improving estimates of viral clearance probabilities will reduce uncertainty in projections of survivor-driven transmission. Long-term follow-up of survivor cohorts, particularly individuals with prolonged viral persistence, remains essential for refining estimates of recrudescence risk and informing post-epidemic surveillance strategies.

### 4.3 Age-dependent viral clearance as the primary driver of survivor-reservoir composition

A consistent finding across all viral-clearance scenarios was the dominant contribution of older male survivors (>35 years) to survivor-driven transmission potential. Older survivors exhibited the longest effective persistence durations and contributed the largest share of the male recrudescence reproduction number across all clearance scenarios. Previous studies have identified age as an important determinant of viral clearance (Thorson et al., 2021; Dyal et al., 2023). Our analysis extends these observations by quantifying their population-level consequences. Under the slowest clearance scenario, older survivors contributed approximately twice as much to the male recrudescence reproduction number as middle-aged survivors and more than five times as much as younger survivors.

These findings indicate that recrudescence risk is not evenly distributed across survivor populations but is concentrated among older male survivors. Age therefore emerges not only as an individual-level determinant of viral clearance but also as a population-level determinant of survivor-driven transmission. The persistence of this age gradient across all clearance scenarios suggests that age remains an important predictor of the epidemiological contribution of the survivor reservoir, even when assumptions about viral clearance vary.

The implications for survivor-reservoir composition are substantial. Older survivors represented only a fraction of survivors but contributed disproportionately to the effective survivor reservoir because of their longer persistence durations. Previous studies have reported that older survivors are more likely to experience severe disease and prolonged viral persistence (Dyal et al., 2023). This creates a compound risk factor in which the individuals most likely to experience severe infection are also those most likely to contribute to long-term recrudescence risk. These results suggest that older male survivors represent a priority group for long-term follow-up and survivor monitoring programmes.

### 4.4 Female contribution as a critical evidence gap

Female contribution consistently increased estimated recrudescence potential across all viral-clearance scenarios. Under the slowest clearance scenario, maximal female contribution increased the posterior mean *R*_*c*_ above one, whereas under baseline and higher clearance assumptions, transmission remained below the persistence threshold. These findings indicate that female survivors may represent a non-negligible component of the post-epidemic reservoir. However, our assessment of female contribution was necessarily indirect because equivalent longitudinal datasets do not exist for female survivors. While EVD RNA has been detected in vaginal secretions, breast milk, and other body fluids (Rodriguez et al., 1999; Keita et al., 2019; Meek et al., 2025), the duration of persistence, shedding dynamics, and transmission efficiency remain poorly characterised. As a result, female contribution could only be explored through scenario analyses rather than empirical estimation.

### 4.5 Implications for surveillance architecture and age-stratified monitoring

Current post-outbreak Ebola surveillance is largely based on the assumption that re-emergence results from new zoonotic spillover events. Our findings support a broader surveillance framework in which the survivor reservoir is recognised as an additional pathway for Ebola re-emergence. The deterministic outbreak simulations further suggest that surveillance intensity should reflect the scale of the preceding epidemic. Following small outbreaks, survivor-driven outbreak pressure remained below the estimated natural spillover rate. In contrast, outbreaks comparable in size to the 2013–2016 West African epidemic generated a transient period during which survivor-driven outbreaks could temporarily exceed zoonotic spillover. These findings support sustained survivor surveillance following large epidemics, particularly during the first few years after outbreak control.

Current WHO guidance recommends safer sex practices for male survivors for at least 12 months in the absence of confirmed negative semen tests (WHO, 2025). Our results further suggest that survivor follow-up could benefit from risk stratification. Older survivors consistently exhibited longer persistence durations and contributed substantially more to the survivor reservoir than younger survivors across all viral-clearance scenarios. Rather than applying identical follow-up strategies to all survivors, programmes could prioritise testing, counselling, and long-term monitoring for individuals most likely to contribute to survivor-driven transmission. Similar risk-based approaches are widely used in HIV care, where monitoring intensity is adapted according to patient risk profiles rather than applied uniformly (Cohen et al., 2012).

Behavioural and social factors may further influence surveillance effectiveness. Juga et al. (2023) demonstrated that Ebola-related stigma reduces healthcare engagement and participation in survivor programmes. Importantly, older survivors reported greater disengagement than younger survivors (Badio et al., 2026). This creates a notable paradox: the survivor group predicted to contribute most to post-epidemic transmission may also be the least likely to remain engaged in long-term monitoring programmes. Integrating biological persistence with behavioural and social determinants may therefore provide a more comprehensive framework for survivor surveillance than approaches based solely on virological monitoring (Bedson et al., 2021).

### 4.6 Limitations

Some limitations should be acknowledged. First, age-specific clearance estimates were derived from relatively limited persistence data. Existing cohorts remain affected by intermittent sampling, finite follow-up periods, and right-censoring, all of which may contribute to underestimation of long-term persistence (Meek et al., 2025). Second, the analysis was restricted primarily to male survivors because equivalent longitudinal datasets for female genital persistence are not available. Although female contribution was explored through scenario analysis, the absence of empirical female persistence parameters remains an important source of uncertainty. Third, the model was not validated against independent survivor cohorts or post-epidemic outbreak datasets. External validation would strengthen confidence in parameter estimates and improve assessment of model performance across different epidemiological settings. Despite these limitations, this study provides the first age-stratified Bayesian quantification of survivor-driven recrudescence risk based on empirical persistence data. The results identify viral clearance, survivor age, epidemic size, and uncertainty surrounding female contribution as the principal determinants of survivor-driven recrudescence in our framework.

## 5. Conclusion

This study developed an age-structured Bayesian framework to quantify the contribution of Ebola survivors to post-epidemic recrudescence using empirical viral persistence data. By integrating survivor persistence, age-specific viral clearance, and uncertainty in female survivor contribution, we quantified how these factors influence the recrudescence reproduction number and the short-term risk of survivor-driven outbreaks. Four main conclusions emerge. First, survivor-driven transmission represents a plausible pathway for Ebola re-emergence, although sustained transmission is unlikely under typical viral-clearance assumptions. The potential for self-sustaining survivor-driven transmission increased under the slowest viral-clearance scenario and with greater female survivor contribution. Second, outbreak size strongly influenced the short-term epidemiological importance of the survivor reservoir. While survivor-driven outbreak pressure remained below the estimated natural spillover rate following small outbreaks, epidemics comparable in size to the 2013–2016 West African epidemic generated a transient period during which survivor-driven outbreaks could temporarily exceed zoonotic spillover. Third, age-dependent viral clearance strongly shaped the survivor reservoir. Older male survivors consistently exhibited longer effective persistence durations and contributed disproportionately to the recrudescence reproduction number across all viral-clearance scenarios. Finally, uncertainty surrounding female survivor contribution remains an important evidence gap. Although explored through scenario analyses, improved empirical data on viral persistence and transmission among female survivors are needed to better quantify their contribution to post-epidemic transmission.

Overall, our findings support expanding Ebola re-emergence risk assessments beyond repeated zoonotic spillover to include survivor reservoirs as a complementary source of future outbreaks. From a public health perspective, the results support sustained survivor surveillance following large epidemics, age-informed monitoring of higher-risk survivor groups, and continued access to survivor testing, counselling, and follow-up programmes.

Future research should prioritise larger longitudinal survivor cohorts, particularly among women, to improve estimates of viral clearance dynamics and survivor-associated transmission. Incorporating intermittent viral shedding, behavioural heterogeneity, and spatial variation into future models will further refine projections of survivor-driven recrudescence and strengthen preparedness for post-epidemic Ebola re-emergence.

## Data Availability

All data produced in the present study are available upon reasonable request to the authors

https://www.cdc.gov/ebola/outbreaks/

## Authors contribution

**Bruno Enagnon Lokonon**: Writing – original draft, Visualization, Validation, Software, Methodology, Formal analysis, Conceptualization.

**Daniel T. Haydon**: Writing – review & editing, Methodology, Conceptualization, Supervision.

**Caroline Fakas**: Writing – review & editing.

**Bassirou Bonfoh**: Writing – review & editing, Supervision.

## Funding

The authors sincerely thank the African Research Excellence Fund (AREF) for its generous financial support, which contributed to the successful completion of this research. This work was also supported by the DELTAS Africa Initiatives I & II [Afrique One-ASPIRE /DEL-15-008 and Afrique One-REACH /DEL-Del-22-011]. Afrique One is funded by a consortium of donors including Science Foundation for Africa (SFA), the Wellcome Trust [107753/A/15/Z and the UK government (UKAID)

## Conflict of interest

The authors declare that they have no known competing financial interests or personal relationships that could have appeared to influence the work reported in this paper.

## Notes

### Competing Interest Statement

The authors have declared no competing interest.

### Author Declarations

https://www.cdc.gov/ebola/outbreaks/ or in published articles

